# Neighborhood Opportunity and Exercise-Related Cardiac Symptoms in Youth Athletes

**DOI:** 10.1101/2025.10.28.25339024

**Authors:** Douglas Corsi, Avinash Saraiya, Grace Qiu, Imran Masood, Abbas H. Zaidi, Jonathan H. Kim, David Shipon

**Affiliations:** Department of Internal Medicine at Rutgers Robert Wood Johnson University Hospital, New Brunswick, NJ; Department of Internal Medicine at University of Maryland Medical System, Baltimore, MD; Sidney Kimmel Medical College at Thomas Jefferson University, Philadelphia, PA; Department of Cardiology, Children’s Hospital of Los Angeles, Los Angeles, CA; Department of Cardiology Nemours Children’s Health, Wilmington, DE; Emory Clinical Cardiovascular Research Institute, Emory University School of Medicine, Atlanta, GA; Division of Cardiology, Thomas Jefferson University Hospital, Philadelphia, PA

**Keywords:** social determinants of health, youth athletes, cardiac screening, health disparities, preparticipation examination

## Abstract

**Background:** Sudden cardiac arrest (SCA) disproportionately affects youths from lower socioeconomic neighborhoods. The underlying mechanisms for this disparity, particularly in youth athletes, remain unclear. While exercise-related cardiac symptoms serve as vital warning signs for identifying at-risk athletes, no studies have examined the prevalence of these symptoms across different socioeconomic strata. We hypothesized that social determinants of health, quantified by the Child Opportunity Index (COI), a validated multidimensional measure of neighborhood conditions, would be associated with cardiac symptoms identified during preparticipation screening.

**Methods:** This retrospective cross-sectional study analyzed data from the Simon’s Heart HeartBytes National Youth Cardiac Registry, a large preparticipation cardiac screening database. Youth athletes aged ≤17 years who completed standardized cardiovascular screening questionnaires were stratified by neighborhood opportunity level using the COI, a validated multidimensional measure of neighborhood conditions affecting child development. Multivariable logistic regression examined associations between COI quintiles and exercise-related cardiac symptoms, adjusting for demographics and comorbidities.

**Results:** Among 10,000 youth athletes analyzed (median age 14.0 years; 38.8% female; 80.3% White), distribution across COI quintiles was 9.8% very low, 5.7% low, 7.7% moderate, 15.9% high, and 61.0% very high. Exercise-related chest pain and exercise-related fatigue demonstrated a higher prevalence in the lowest COI quintile (p<0.001). After adjustment for age, sex, race, physical activity, and comorbidities, participants from the very high COI quintile had significantly lower odds of exercise-related chest pain (p<0.001) and exercise-related fatigue (p=0.004) compared with the very low COI quintile.

**Conclusions:** Youths from lower-opportunity neighborhoods were underrepresented in screening yet showed higher prevalence of exercise-related symptoms. Whether cardiac or noncardiac in origin, these disparities burden under-resourced communities with increased evaluations. Addressing screening inequities and understanding symptom patterns across socioeconomic strata are critical for equitable cardiovascular care. Prospective studies are needed to determine the clinical significance of these symptom patterns and develop equitable screening strategies.

## Introduction

Out-of-hospital sudden cardiac arrest (SCA) affects more than 20,000 children annually in the United States, representing a devastating clinical event with profound implications for pediatric populations^1,2^. The incidence and outcomes of SCA demonstrate significant associations with social determinants of health (SDOH), with evidence indicating higher rates of pediatric out-of-hospital cardiac arrest in neighborhoods characterized by lower socioeconomic status^3–5^. These disparities manifest through reduced rates of bystander cardiopulmonary resuscitation (CPR), utilization of automated external defibrillators (AEDs), and return of spontaneous circulation in vulnerable communities^4^.

Preparticipation cardiac screening represents a critical intervention for potentially identifying young athletes with cardiovascular conditions (such as hypertrophic cardiomyopathy (HCM), anomalous coronary artery origin, and Long QT syndrome (LQTS)) that are associated with increased risk of sudden cardiac arrest/death (SCA/SCD). The American Heart Association (AHA) and the American College of Cardiology (ACC) recommend preparticipation cardiovascular screening for athletes, which begins with a targeted cardiovascular medical and family history, as well as a physical examination^6,7^. Cardiac symptom assessment is central to risk stratification, as exercise-related symptoms are established red flags for conditions associated with SCA/SCD risk.^7–11^ However, the sensitivity of symptom assessment for detecting cardiovascular pathology remains limited^7^. Indeed, over one-third of pediatric patients who ultimately receive such diagnoses initially present with concerning symptoms^12^. Mounting evidence demonstrates that SDOH are closely linked to inequities in SCA prevention, risk, and care^4,13,14^. This is particularly apparent in Black communities, where athletes experiencing SCA disproportionately originate from neighborhoods with greater socioeconomic deprivation^15^. These disparities underscore the urgent need to systematically examine how neighborhood-level factors influence the identification of SCA risk in youth athletes.

To address this knowledge gap, we utilized the Child Opportunity Index (COI) 3.0, a multidimensional measure that incorporates 29 indicators across educational, health/environmental, and socioeconomic domains, comprehensively capturing neighborhood-level conditions^16^. The COI has demonstrated utility in examining SDOH associations with various pediatric health outcomes, providing a granular assessment beyond traditional socioeconomic metrics. This retrospective analysis examined the association between COI and cardiac symptoms that serve as red flags for cardiovascular conditions associated with increased SCA/SCD risk, as identified during preparticipation screening in a national registry of youth athletes.

## Methods

### Study Design and Population

This retrospective cross-sectional study utilized the HeartBytes National Youth Cardiac Registry, maintained by Simon’s Heart—a nonprofit organization focused on SCD prevention in children and young adults. Data were collected from participants attending voluntary cardiac screening events conducted nationwide between April 2015 and May 2024. Participant information was recorded using comprehensive questionnaires that encompassed medical history, family history, and symptom assessment. Screening questionnaires were based on the AHA’s 14-element screening recommendation^8^, modified for the HeartBytes registry as previously described^17^ (Figure 1). The study protocol received approval from the Institutional Review Board. The initial cohort consisted of 11,430 individuals who completed comprehensive medical and family history questionnaires during their preparticipation evaluation. Inclusion criteria were: (1) age 17 years or younger at time of screening, (2) complete demographic and zip code data, and (3) valid residential zip code data enabling COI mapping.

**Figure 1:**
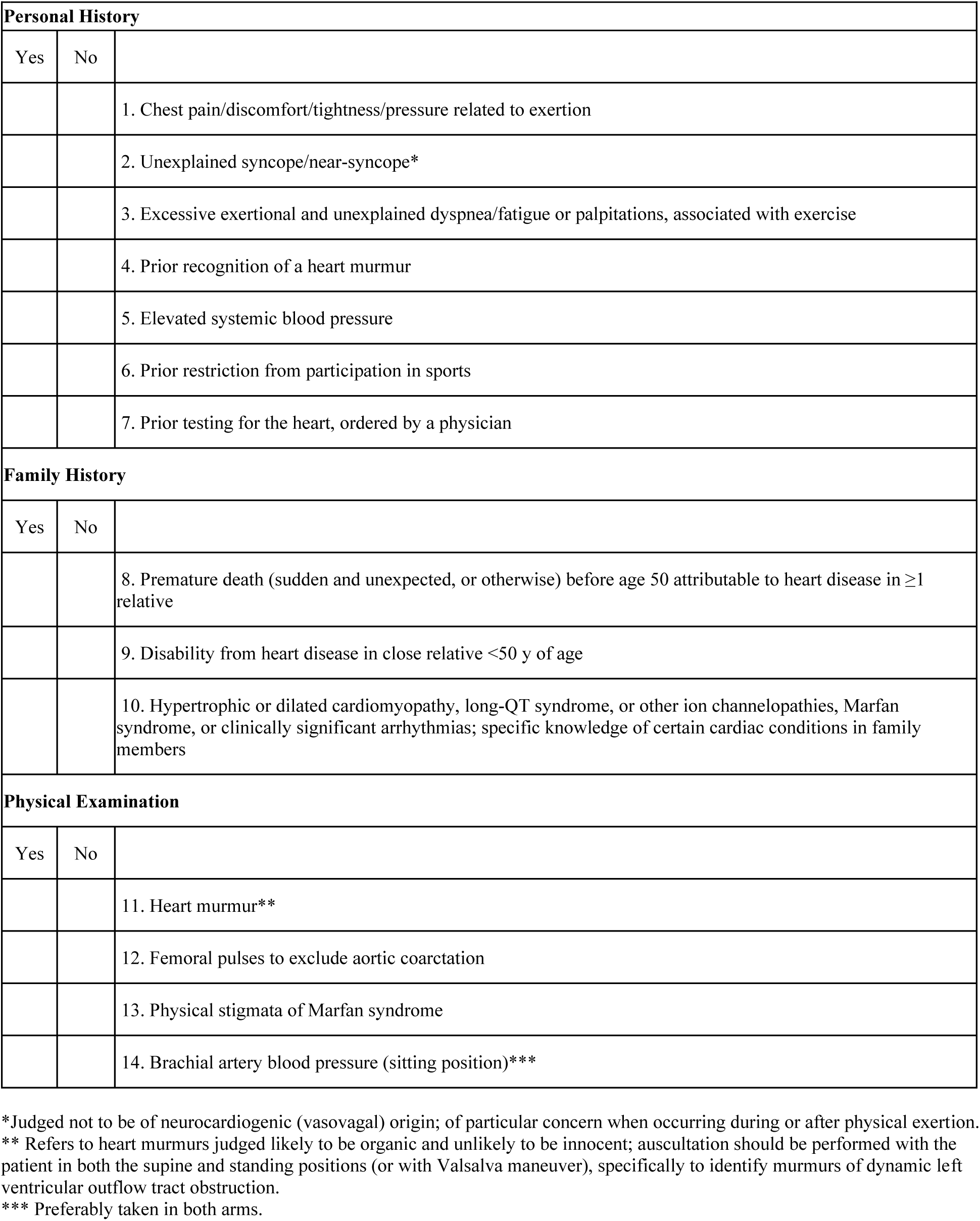
The 14-Element AHA Cardiovascular Screening Checklist for Congenital and Genetic Heart Disease^15^

### Variables of Interest

#### Demographic and Physical Activity

Demographic variables included age at screening, sex, and race and/or ethnicity (White, Black, Asian/Pacific Islander, Hispanic, African, Native American, and Other). Physical activity levels were categorized into four groups: <2 hours/week, 2-5 hours/week, 5-10 hours/week, and >10 hours/week.

#### Cardiac Symptoms and History

Cardiac symptom assessment included history of exercise-related symptoms (chest pain during exercise, syncope after exercise, difficulty breathing during exercise, easily tired during exercise, heart racing without apparent cause). Cardiovascular history variables included hypertension, hypercholesterolemia, Kawasaki disease, heart infection, and heart murmur.

#### Comorbidities

Comorbidities included asthma, anemia, sickle cell anemia, diabetes mellitus, kidney disease, hearing problems, attention-deficit/hyperactivity disorder (ADHD), and anxiety/depression. Hearing problems were included given the established association between congenital hearing loss and long QT syndrome (LQTS), particularly Jervell and Lange-Nielsen syndrome^9,18^.

Follow-up data regarding the completion of cardiac evaluation among participants with positive screening findings were not available in this cross-sectional analysis.

### Child Opportunity Index

The primary exposure variable was neighborhood opportunity level, measured using the COI 3.0. The COI provides a comprehensive multidimensional measure of neighborhood conditions affecting child development through 29 indicators across three domains: educational opportunity, health and environmental opportunity, and social and economic opportunity^16^. Participants’ residential zip codes were geocoded and mapped to corresponding census tracts to obtain associated COI scores. Scores were categorized into national quintiles: very low (≤20th percentile), low (21st-40th percentile), moderate (41st-60th percentile), high (61st-80th percentile), and very high (>80th percentile). Secondary analyses examined the three component domains independently.

### Statistical Analysis

Descriptive statistics were calculated to characterize demographic and clinical distributions across COI levels. Continuous variables were reported as medians with interquartile ranges (IQRs) due to non-normal distributions. Categorical variables were compared using Pearson’s chi-square test. Primary outcomes included exercise-related cardiac symptoms and cardiovascular history findings. Secondary outcomes encompassed exercise patterns and participation levels.

Multivariable logistic regression models were used to evaluate the associations between COI level and cardiac symptoms. Initial models adjusted for age, sex/gender, and race/ethnicity. Fully adjusted models incorporated comprehensive covariate adjustment, including age at screening, sex, race, physical activity, and all assessed comorbidities (asthma, anemia, sickle cell anemia, diabetes, kidney disease, hearing problems, hypertension, ADHD, and anxiety/depression). These models assessed the robustness of associations after accounting for potential clinical confounders that may mediate relationships between neighborhood opportunity and cardiac symptoms.

Odds ratios (OR) with 95% confidence intervals (CI) were calculated using the very low COI level as the reference category. Model discrimination was assessed using the C statistic. Statistical significance was defined as p<0.05. Missing data were not imputed, and complete case analysis was performed. All analyses were conducted using IBM SPSS Version 29.0.2.0 and R statistical software version 4.1.0.

## Results

### Study Population Characteristics

The study included 10,000 youth athletes with a median age of 14 years (IQR 3) and a median COI of 85 (IQR 27), of whom 38.8% were female and 80.3% were White (Table 1). Distribution across COI quintiles ranged from 9.8% very low COI to 61.0% very high COI (Figure 2). Racial composition differed across COI quintiles (p<0.001), with Black participants comprising 31.0% of the very low COI quintile compared with 6.4% of the very high COI quintile (Figure 3). Physical activity levels did not differ significantly across COI quintiles (p = 0.111), with similar proportions of individuals engaging in high-intensity exercise (>10 hours/week) across all quintiles (Table 1).

**Figure 2:**
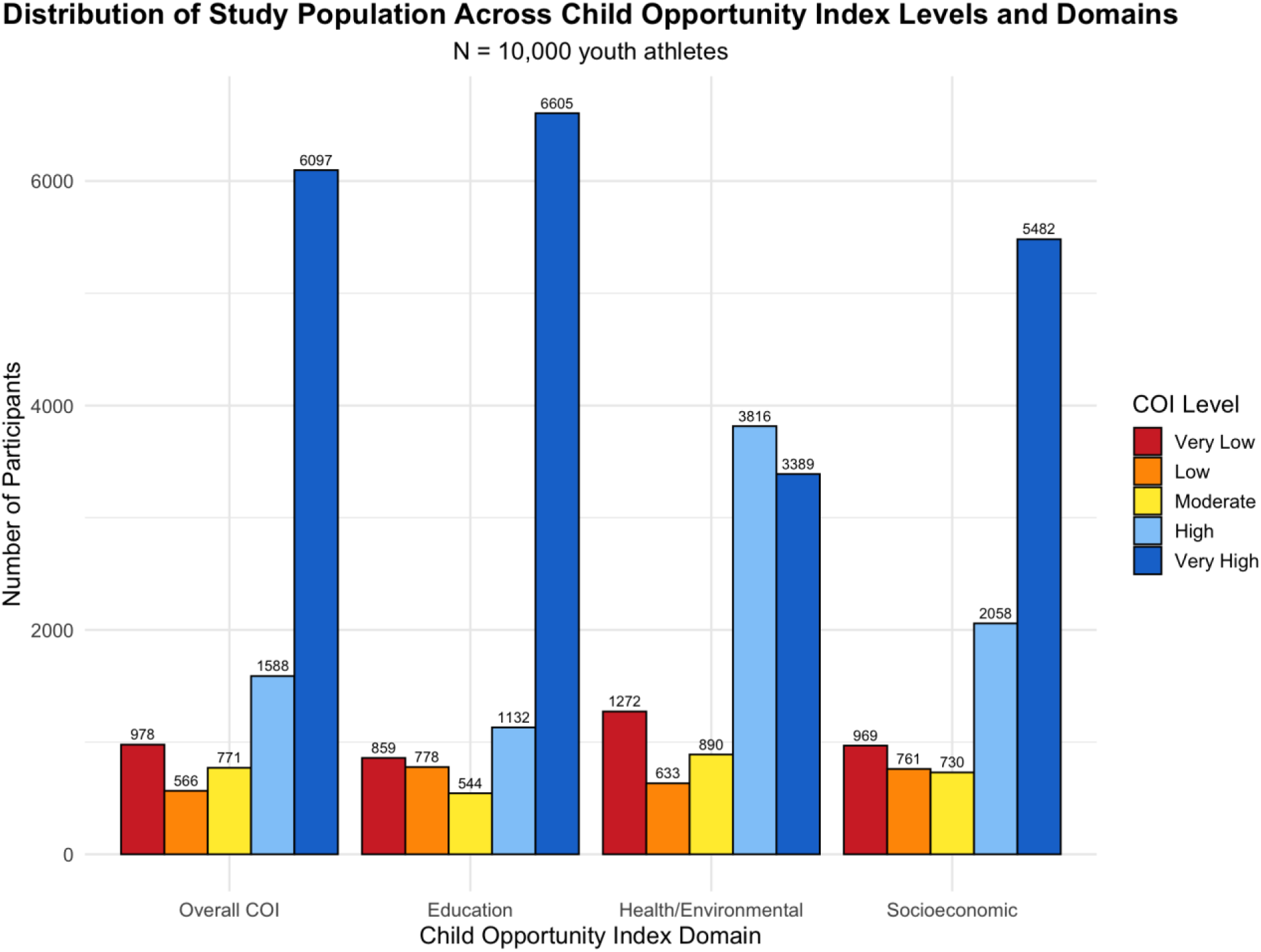
Distribution of Study Population Across COI and COI Domain Quintiles

**Figure 3:**
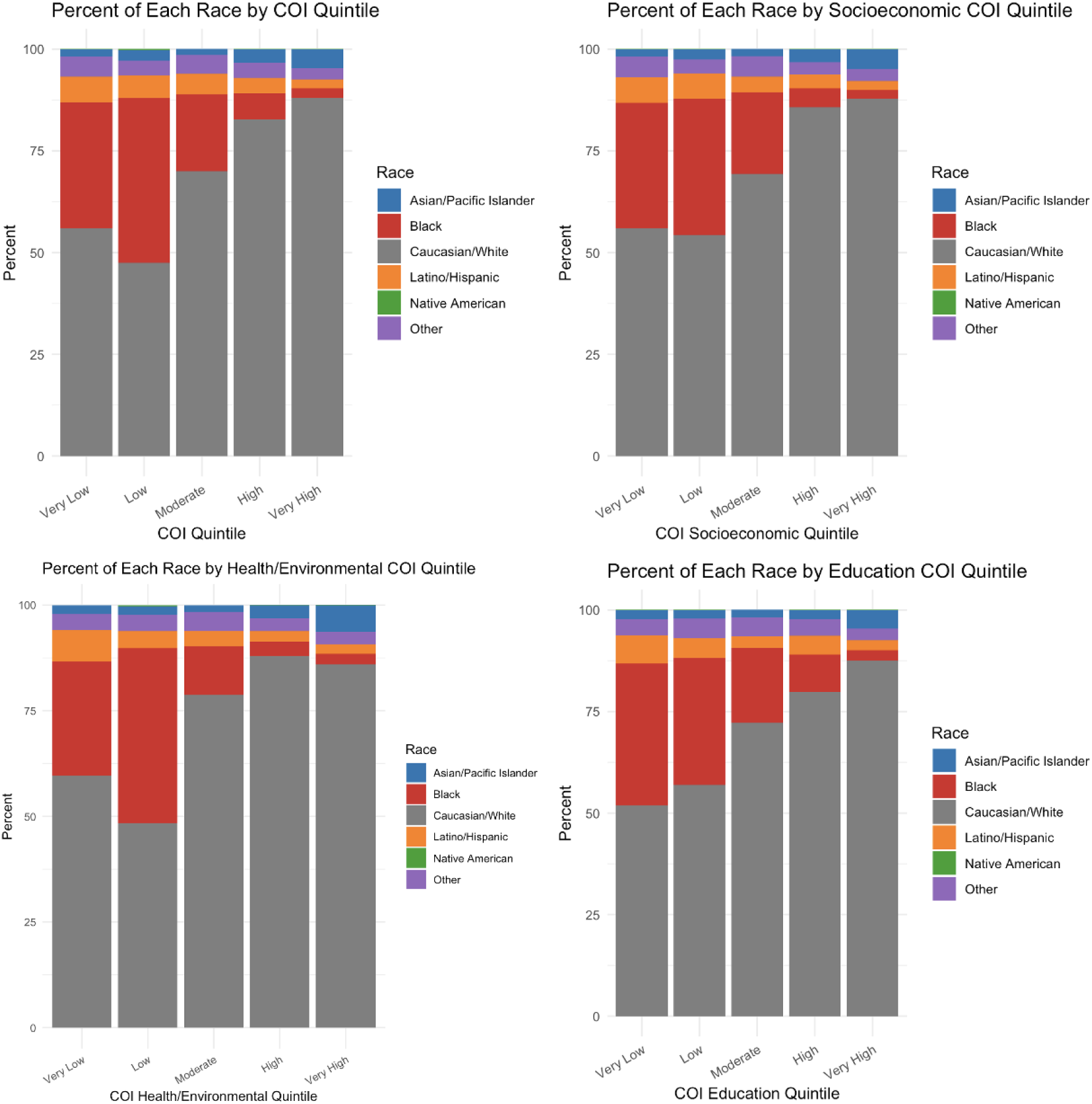
Distribution of Race Across Childhood Opportunity Index (COI) and COI Domain

**Table 1:** Population Characteristics.

| **Variable / Category** | **Very Low**<br>**(978 / 9.8%)** | **Low**<br>**(566 / 5.7%)** | **Moderate**<br>**(771 / 7.7%)** | **High**<br>**(1588 / 15.9%)** | **Very High**<br>**(6097 / 61.0%)** | **Total** | **p-value** |
| --- | --- | --- | --- | --- | --- | --- | --- |
| Age, median (IQR) | 14 (3) | 14 (3) | 14 (2) | 14 (3) | 14 (3) | 14 (3) | <0.001 |
| *Gender* |  |  |  |  |  |  | <0.001 |
| Female | 335 (34.2%) | 2479 (40.7%) | 279 (36.2%) | 185 (32.7%) | 607 (38.2%) | 3885<br>(38.8%) |  |
| *Race* |  |  |  |  |  |  | <0.001 |
| White | 547 (55.9%) | 5364 (88.0%) | 540 (95.4%) | 269 (47.6%) | 1314 (82.8%) | 8034<br>(80.3%) |  |
| Black | 303 (31.0%) | 142 (2.3%) | 145 (25.6%) | 229 (40.5%) | 101 (6.4%) | 920 (9.2%) |  |
| Asian/Pacific<br>Islander | 17 (1.7%) | 280 (4.6%) | 11 (1.9%) | 15 (2.7%) | 53 (3.3%) | 376 (3.8%) |  |
| Hispanic | 62 (6.3%) | 135 (2.2%) | 39 (6.9%) | 31 (5.5%) | 61 (3.8%) | 328 (3.3%) |  |
| Native American | 1 (0.1%) | 5 (0.1%) | 0 (0.0%) | 1 (0.2%) | 1 (0.1%) | 8 (0.1%) |  |
| Other | 48 (4.9%) | 171 (2.8%) | 36 (6.4%) | 21 (3.7%) | 58 (3.7%) | 334 (3.3%) |  |
| *Physical Activity* |  |  |  |  |  |  | 0.111 |
| 2–5 hrs/week | 183 (18.7%) | 1117 (18.3%) | 148 (26.2%) | 108 (19.1%) | 302 (19.0%) | 1858<br>(18.6%) |  |
| 5–10 hrs/week | 356 (36.4%) | 2312 (37.9%) | 280 (49.5%) | 198 (35.0%) | 534 (33.6%) | 3680<br>(36.8%) |  |
| <2 hrs/week | 46 (4.7%) | 294 (4.8%) | 31 (5.5%) | 38 (6.7%) | 72 (4.5%) | 481 (4.8%) |  |
| >10 hrs/week | 393 (40.2%) | 2374 (38.9%) | 312 (55.1%) | 222 (39.2%) | 680 (42.8%) | 3981<br>(39.8%) |  |
| Comorbidities |  |  |  |  |  |  |  |
| Asthma | 233 (23.8%) | 127 (22.4%) | 169 (21.9%) | 286 (18.0%) | 991 (16.3%) | 1806<br>(18.1%) | <.001 |
| Anemia | 10 (1.0%) | 9 (1.6%) | 15 (1.9%) | 14 (0.9%) | 48 (0.8%) | 96 (1.0%) | 0.015 |
| Sickle Cell Anemia | 2 (0.2%) | 4 (0.7%) | 2 (0.3%) | 0 (0.0%) | 1 (0.0%) | 9 (0.1%) | <.001 |
| Diabetes | 2 (0.2%) | 1 (0.2%) | 6 (0.8%) | 1 (0.1%) | 16 (0.3%) | 26 (0.3%) | 0.031 |
| Kidney Disease | 1 (0.1%) | 0 (0.0%) | 0 (0.0%) | 3 (0.2%) | 5 (0.1%) | 9 (0.1%) | 0.559 |
| Hearing Problems | 3 (0.3%) | 4 (0.7%) | 11 (1.4%) | 10 (0.6%) | 36 (0.6%) | 64 (0.6%) | 0.05 |
| High Blood Pressure | 9 (0.9%) | 3 (0.5%) | 1 (0.1%) | 5 (0.3%) | 7 (0.1%) | 25 (0.3%) | <.001 |
| ADHD | 82 (8.4%) | 43 (7.6%) | 61 (7.9%) | 126 (7.9%) | 448 (7.3%) | 760 (7.6%) | 0.778 |
| Anxiety/Depression | 28 (2.9%) | 18 (3.2%) | 28 (3.6%) | 86 (5.4%) | 337 (5.5%) | 497 (5.0%) | <.001 |

### Exercise-Related Symptoms and Screening History

Exercise-related symptoms varied significantly across COI quintiles (Table 2, Figure 4). Four symptoms demonstrated significant variation across COI quintiles: chest pain during exercise, feeling easily tired during exercise (both p < 0.001), difficulty breathing during exercise (p = 0.013), and heart racing without apparent cause (p = 0.022). Among cardiovascular history variables, only previous heart murmur showed significant differences across quintiles (p<0.001).

**Figure 4:**
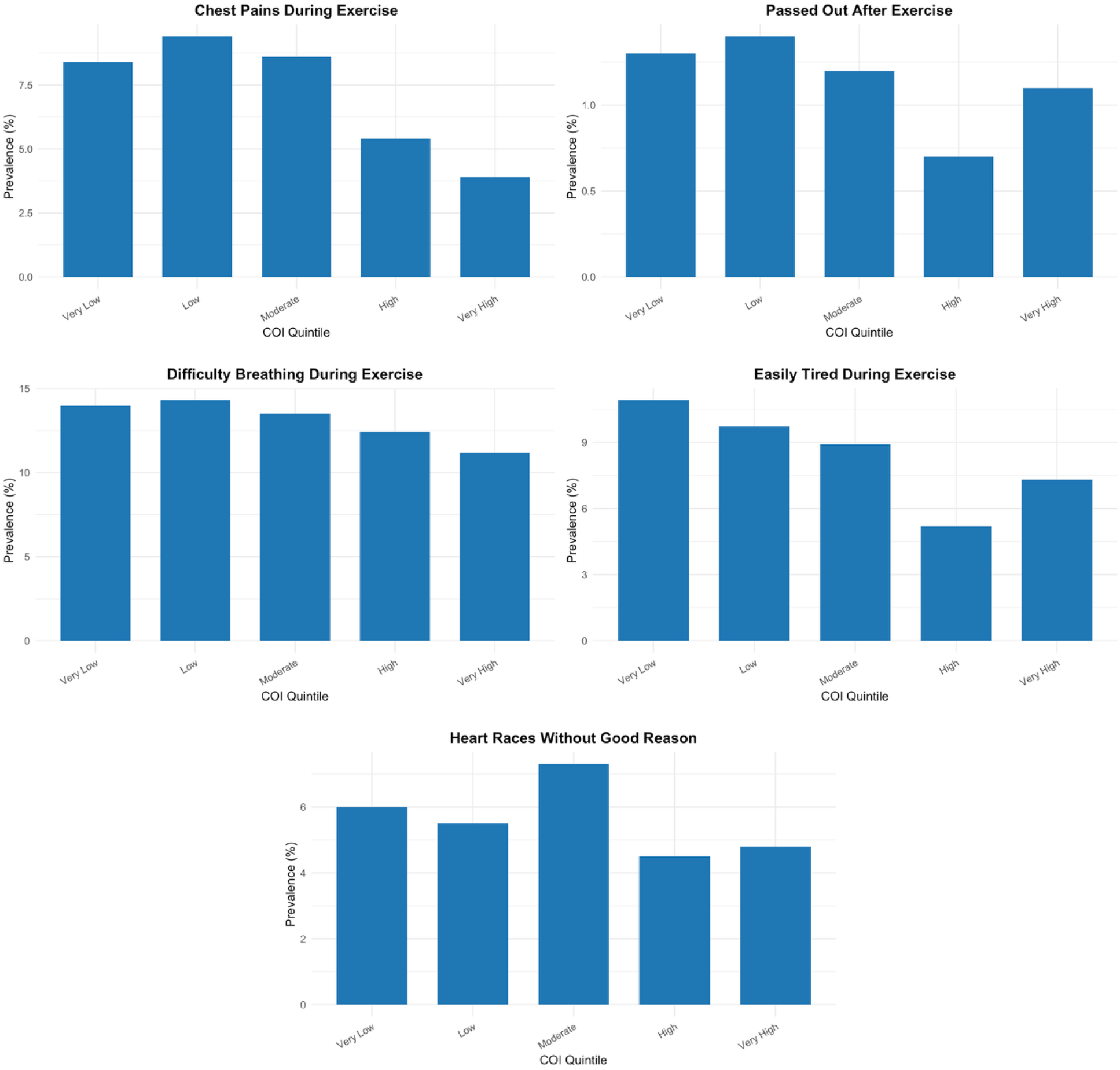
Prevalence of Exercise-Related Cardiac Symptoms by Child Opportunity Index Level

**Table 2:** Prevalence of Exercise-Related Cardiac Symptoms and Cardiovascular History by Child Opportunity Index Level.

| Condition | **Very Low** | **Low** | **Moderate** | **High** | **Very High** | **Total** | p-value |
| --- | --- | --- | --- | --- | --- | --- | --- |
| *Exercise-Related Symptoms* |  |  |  |  |  |  |  |
| Chest Pains During Exercise | 82 (8.4%) | 53 (9.4%) | 66 (8.6%) | 86 (5.4%) | 235 (3.9%) | 522 (5.2%) | <.001 |
| Passed Out After Exercise | 13 (1.3%) | 8 (1.4%) | 9 (1.2%) | 11 (0.7%) | 68 (1.1%) | 109 (1.1%) | 0.482 |
| Difficulty Breathing During Exercise | 137 (14.0%) | 81 (14.3%) | 104 (13.5%) | 197 (12.4%) | 680 (11.2%) | 1199 (12.0%) | 0.013 |
| Easily Tired During Exercise | 107 (10.9%) | 55 (9.7%) | 69 (8.9%) | 98 (6.2%) | 446 (7.3%) | 775 (7.8%) | <.001 |
| Heart Races Without Good Reason | 59 (6.0%) | 31 (5.5%) | 56 (7.3%) | 71 (4.5%) | 295 (4.8%) | 512 (5.1%) | 0.022 |
| *Cardiovascular History* |  |  |  |  |  |  |  |
| Previous High Blood Pressure | 7 (0.7%) | 3 (0.5%) | 2 (0.3%) | 5 (0.3%) | 9 (0.1%) | 26 (0.3%) | 0.014 |
| Previous High Cholesterol | 13 (1.3%) | 13 (2.3%) | 11 (1.4%) | 35 (2.2%) | 99 (1.6%) | 171 (1.7%) | 0.293 |
| Previous Kawasaki Disease | 1 (0.1%) | 0 (0.0%) | 1 (0.1%) | 0 (0.0%) | 11 (0.2%) | 13 (0.1%) | 0.399 |
| Previous Heart Infection | 2 (0.2%) | 0 (0.0%) | 0 (0.0%) | 0 (0.0%) | 3 (0.0%) | 5 (0.1%) | 0.189 |
| Previous Heart Murmur Diagnosis* | 67 (6.9%) | 38 (6.7%) | 40 (5.2%) | 89 (5.6%) | 223 (3.7%) | 457 (4.6%) | <.001 |

\*All cardiovascular history assessment specifically captured whether participants had ever been informed by a healthcare provider of these diagnoses, thus representing participant recollection of previous clinical diagnosis rather than objective findings at screening.

### Comorbidities

Multiple comorbidities demonstrated significant variation across COI quintiles. Asthma prevalence was highest in the very low COI quintile (23.8%) compared with the very high COI quintile (16.3%) (p <0.001). Anemia showed similar patterns (1.0% vs. 0.8%, p = 0.015). Sickle cell anemia was found exclusively in very low, low, and moderate COI quintiles (p<0.001). Hypertension prevalence was significantly higher in lower COI quintiles (p<0.001). Anxiety/depression demonstrated the highest prevalence in high and very high COI quintiles (p<0.001).

### Multivariable Analysis

After comprehensive adjustment, participants from the very high COI quintile demonstrated significantly lower odds of exercise-related chest pain (aOR 0.52, 95% CI 0.39-0.69, p<0.001) and exercise-related fatigue (aOR 0.70, 95% CI 0.54-0.89, p=0.004) compared with the very low COI quintile (Table 3, Figure 5). For exercise-related chest pain, high (aOR 0.71, p = 0.040) and very high (aOR 0.52, p < 0.001) COI quintiles had significantly lower odds compared to the very low COI quintile. Easily tired during exercise also showed persistent associations, with high (aOR 0.57, p < 0.001) and very high (aOR 0.70, p = 0.004) COI quintiles demonstrating significantly lower odds. Post-exercise syncope and difficulty breathing during exercise showed no statistically significant associations with COI levels after full adjustment for confounding factors.

**Figure 5:**
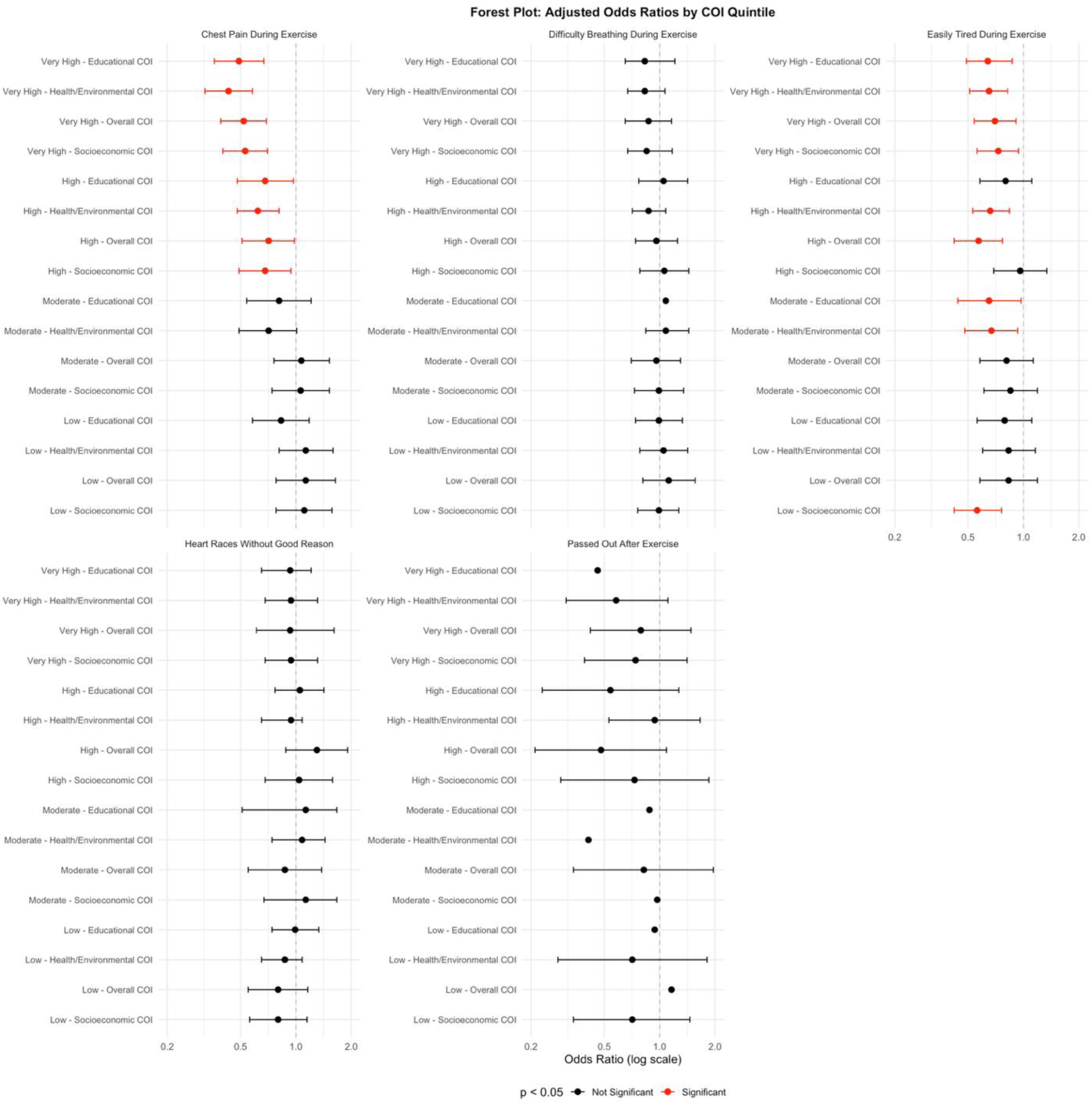
Child Opportunity Index and Exercise-Related Cardiac Symptoms. Multivariable adjusted odds ratios (95% CI) for significant associations between Child Opportunity Index levels and exercise-related cardiac symptoms. Adjusted for age at screening, gender, race, and all assessed comorbidities (asthma, anemia, sickle cell anemia, diabetes, kidney disease, hearing problems, hypertension, ADHD, and anxiety/depression Reference: Very Low COI. COI, Child Opportunity Index

**Table 3:**
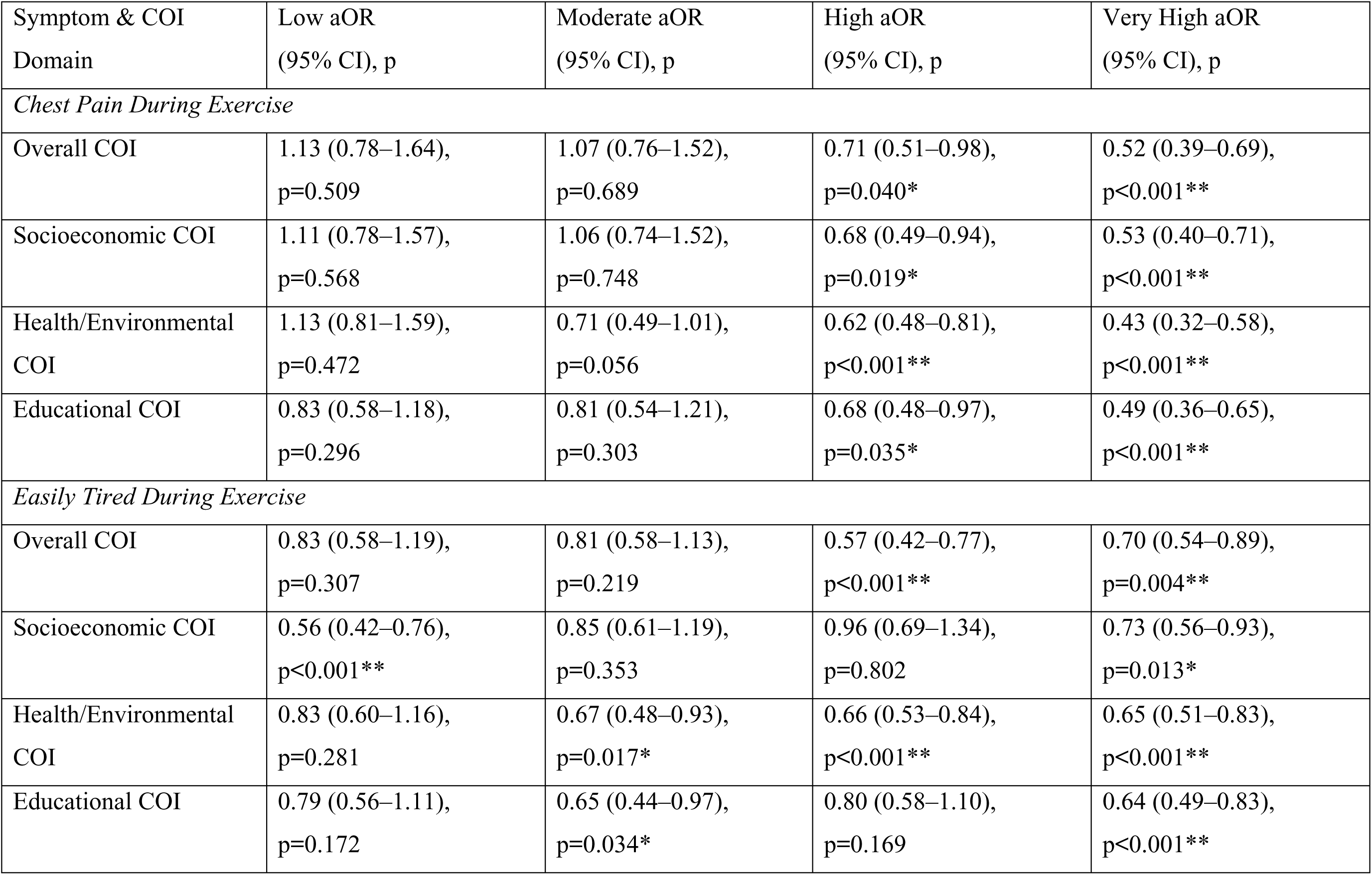
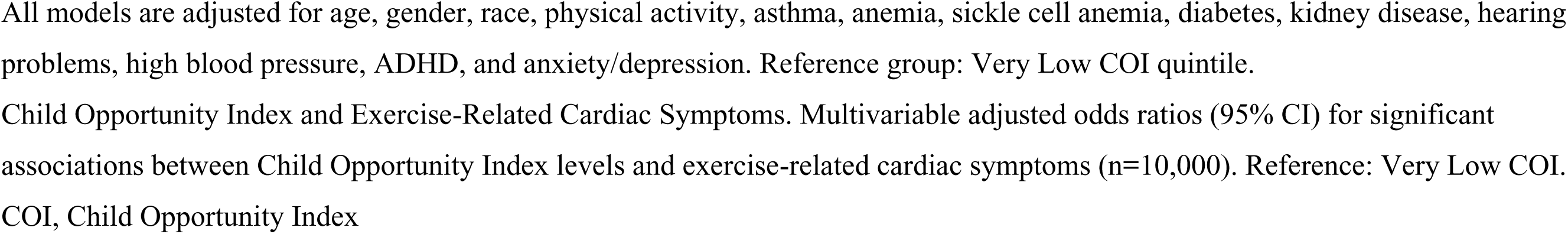
Adjusted Odds Ratios (aOR) for Cardiac Screening Outcomes by COI Quintile.

| Symptom & COI Domain | Low aOR<br>(95% CI), p | Moderate aOR<br>(95% CI), p | High aOR<br>(95% CI), p | Very High aOR<br>(95% CI), p |
| --- | --- | --- | --- | --- |
| *Chest Pain During Exercise* |  |  |  |  |
| Overall COI | 1.13 (0.78–1.64),<br>p=0.509 | 1.07 (0.76–1.52),<br>p=0.689 | 0.71 (0.51–0.98),<br>p=0.040* | 0.52 (0.39–0.69),<br>p<0.001** |
| Socioeconomic COI | 1.11 (0.78–1.57),<br>p=0.568 | 1.06 (0.74–1.52),<br>p=0.748 | 0.68 (0.49–0.94),<br>p=0.019* | 0.53 (0.40–0.71),<br>p<0.001** |
| Health/Environmental COI | 1.13 (0.81–1.59),<br>p=0.472 | 0.71 (0.49–1.01),<br>p=0.056 | 0.62 (0.48–0.81),<br>p<0.001** | 0.43 (0.32–0.58),<br>p<0.001** |
| Educational COI | 0.83 (0.58–1.18),<br>p=0.296 | 0.81 (0.54–1.21),<br>p=0.303 | 0.68 (0.48–0.97),<br>p=0.035* | 0.49 (0.36–0.65),<br>p<0.001** |
| *Easily Tired During Exercise* |  |  |  |  |
| Overall COI | 0.83 (0.58–1.19),<br>p=0.307 | 0.81 (0.58–1.13),<br>p=0.219 | 0.57 (0.42–0.77),<br>p<0.001** | 0.70 (0.54–0.89),<br>p=0.004** |
| Socioeconomic COI | 0.56 (0.42–0.76),<br>p<0.001** | 0.85 (0.61–1.19),<br>p=0.353 | 0.96 (0.69–1.34),<br>p=0.802 | 0.73 (0.56–0.93),<br>p=0.013* |
| Health/Environmental COI | 0.83 (0.60–1.16),<br>p=0.281 | 0.67 (0.48–0.93),<br>p=0.017* | 0.66 (0.53–0.84),<br>p<0.001** | 0.65 (0.51–0.83),<br>p<0.001** |
| Educational COI | 0.79 (0.56–1.11),<br>p=0.172 | 0.65 (0.44–0.97),<br>p=0.034* | 0.80 (0.58–1.10),<br>p=0.169 | 0.64 (0.49–0.83),<br>p<0.001** |

All models are adjusted for age, gender, race, physical activity, asthma, anemia, sickle cell anemia, diabetes, kidney disease, hearing problems, high blood pressure, ADHD, and anxiety/depression. Reference group: Very Low COI quintile.
Child Opportunity Index and Exercise-Related Cardiac Symptoms. Multivariable adjusted odds ratios (95% CI) for significant associations between Child Opportunity Index levels and exercise-related cardiac symptoms (n=10,000). Reference: Very Low COI. COI, Child Opportunity Index

### COI Domain-Specific Analysis

Domain-specific analysis of the COI demonstrated distinct patterns across component domains. For exercise-related chest pain, the health/environmental domain exhibited the most robust associations, with moderate, high, and very high quintiles showing significantly lower adjusted odds compared with very low (aOR range: 0.41–0.65, all p≤0.018). Similarly, higher socioeconomic opportunity was associated with decreased odds in high and very high quintiles (aOR 0.67 and 0.50, respectively; both p≤0.014), while educational opportunity was associated with reduced odds in low and very high quintiles (aOR 0.65 and 0.47; both p≤0.014). For exercise-related fatigue, higher opportunity levels in the health/environmental, socioeconomic, and educational domains were consistently associated with significantly lower odds across multiple quintiles, with the health/environmental domain demonstrating the most robust and consistent associations (Table 3).

## Discussion

This study demonstrates significant associations between neighborhood opportunity levels and the prevalence of exercise-related cardiac symptoms reported during preparticipation screening in youth athletes. Youths from the lowest COI quintile reported higher rates of exercise-related chest pain and fatigue compared with those from higher opportunity neighborhoods, with associations persisting after comprehensive adjustment for demographics and comorbidities. Additionally, we identified underrepresentation of lower-opportunity neighborhoods in screening participation, with only 15.5% of participants from the lowest two COI quintiles compared to 61.0% from the highest quintile. While previous studies have documented associations between SDOH and SCA incidence^3^ and post-arrest outcome^4,5^, our findings uniquely examine symptom prevalence during routine preparticipation screening— before any cardiac events or diagnoses occur. These findings raise important questions about the underlying etiology and clinical significance of these symptom disparities, as well as the issue of healthcare equity in preparticipation screening.

The distribution of participants across COI quintiles—15.5% from very low and low opportunity areas compared with 61.0% from very high—likely reflects documented patterns of healthcare access inequity. Schools and athletic programs in higher-opportunity neighborhoods typically possess greater administrative infrastructure, financial resources, and parental advocacy networks necessary to organize screening events and establish referral pathways to pediatric cardiology when screening identifies concerning findings^7,19,20^. Conversely, institutions serving lower opportunity populations face competing priorities that often preclude them from providing comprehensive cardiovascular evaluation of at-risk athletes^19,20^. These access imbalances parallel broader patterns of preventive care, wherein populations with the highest cardiovascular risk encounter the most significant barriers to early detection and intervention^19,20^. Despite this marked underrepresentation of lower-opportunity neighborhoods, the higher symptom prevalence identified among those who did access screening warrants careful examination of its clinical significance and potential etiologies.

The observed differences in symptom prevalence may have multiple explanations, including underlying cardiovascular pathology and noncardiac etiologies. While exercise-related chest pain and fatigue require assessment for numerous underlying structural and arrhythmogenic disorders^7–11,21^, it is essential to note that among young competitive athletes, only approximately 6% of exercise-related chest pain stems from underlying cardiovascular disease^22^. Without follow-up cardiovascular evaluation data or clinical outcomes in this cohort, we cannot determine whether the higher symptom prevalence in lower COI neighborhoods reflects actual differences in cardiovascular pathology, environmental factors affecting exercise tolerance, psychosocial influences on symptom reporting, or variations in health literacy and symptom interpretation. Nevertheless, the clinical relevance of exercise-related symptoms in identifying at-risk youths remains an important consideration. In pediatric patients with confirmed cardiac diagnoses, symptomatic presentations constitute the initial referral reason in over one-third of cases, with exercise-related chest pain and exercise intolerance commonly reported as the presenting symptoms in LQTS, HCM, and ALCA-R^12,23–25^, though many cases are detected incidentally. Furthermore, among young athletes with established HCM, a single-center analysis revealed that 7% reported exertional symptoms^26^. Given the multifactorial nature of symptom reporting and the significant associations observed in our study, examining the specific COI domains provides valuable insights into potential underlying mechanisms. The health/environmental and socioeconomic COI domains demonstrated substantial associations with cardiac symptoms. Environmental factors disproportionately affecting lower-opportunity neighborhoods—including air pollution, excessive heat exposure, and poor air quality—can precipitate exercise-related symptoms through both cardiac and non-cardiac pathways, such as oxidative stress, endothelial dysfunction, and inflammatory cascade activation^27^. These same environmental factors also precipitate non-cardiac symptoms through respiratory irritation and bronchial hyperresponsiveness. For example, exercise-induced bronchospasm increases significantly in urban environments with PM2.5 levels exceeding 12 μg/m³ due to pro-inflammatory and airway-irritant effects^28,29^. Additionally, psychosocial factors may influence symptom perception and reporting. Youths from lower socioeconomic backgrounds demonstrate higher rates of anxiety disorders and of somatoform presentations, both of which frequently manifest as exercise-related cardiopulmonary symptoms^30,31^. Furthermore, health literacy considerations may play an important role. Limited health literacy—more prevalent among individuals with lower socioeconomic status and those with less education—may affect how youth and their families recognize and report warning symptoms, such as exertional chest pain, syncope, or palpitations, as well as their understanding of the importance of seeking timely medical evaluation^32,33^. These multiple potential etiologies underscore the complexity of interpreting symptom data without corresponding diagnostic outcomes.

Our findings support existing recommendations from the American Heart Association and the American College of Cardiology, emphasizing the need for equitable implementation of preparticipation screening across all communities^6,7^. The data reinforce several established principles while highlighting persistent implementation challenges. First, communities require adequate infrastructure and resources to conduct screening appropriately if it is to be implemented effectively. This includes access to clinicians with expertise in athlete-specific ECG interpretation, established referral pathways to pediatric cardiology, and diagnostic capabilities for secondary evaluation when initial screening raises concerns^8,34^. Without these foundational elements, screening programs risk generating false positives that lead to unnecessary testing, unwarranted disqualification, and psychological distress. Second, the stark underrepresentation of lower opportunity neighborhoods in our cohort, combined with their higher symptom prevalence, suggests that current passive recruitment models may not reach populations with potentially higher cardiovascular risk. If screening programs are implemented, consideration should be given to active outreach strategies that reduce structural barriers to participation across all communities. Third, the higher symptom prevalence observed in lower opportunity neighborhoods—regardless of the underlying etiology—indicates that these communities may face increased downstream healthcare utilization due to symptom-triggered evaluations. This places an additional burden on populations already facing significant resource constraints. Strategic placement of AEDs and community-based CPR training programs may help address documented disparities in cardiac arrest outcomes, as lower socioeconomic areas demonstrate reduced rates of bystander CPR and AED utilization^4,35,36^. The multidisciplinary approach described by van Hattum et al.^37^ demonstrates how coordinated care can improve diagnostic accuracy while minimizing unnecessary restrictions. Addressing transportation barriers, appointment accessibility, and healthcare navigation—particularly for families from lower opportunity neighborhoods—may help ensure appropriate follow-up when indicated.

### Limitations

Our study has several significant limitations. Most critically, we lack follow-up data on cardiovascular evaluations or clinical outcomes for participants with positive screening findings. This prevents conclusions about whether higher symptom rates in lower COI neighborhoods indicate increased cardiovascular disease burden. Second, the HeartBytes Registry employs community-initiated recruitment, with screening events organized by individual schools and athletic programs rather than a systematic outreach approach. This methodology likely contributed to the overrepresentation of higher-opportunity neighborhoods in our sample, as these communities may have a greater capacity to coordinate screening logistics. Selection bias is likely present, as our sample was limited to those who attended voluntary screening events. Third, the data collection methodology employed a non-standard categorization of race and ethnicity, capturing only racial categories without a separate ethnicity designation. This methodological constraint prevented disaggregation of ethnicity from racial categories, potentially obscuring important heterogeneity within racial groups. Fourth, the self-reported nature of symptom data introduces potential reporting bias, which may vary systematically across socioeconomic groups. Finally, while our models adjusted for asthma and anxiety/depression, statistical adjustment has inherent limitations in disentangling the complex relationships between neighborhood factors, comorbid conditions, and symptom manifestation.

## Conclusion

Youth athletes from lower-opportunity neighborhoods were significantly underrepresented in preparticipation screening programs and demonstrated higher prevalence of exercise-related symptoms when screened. While these symptoms represent established indicators requiring cardiovascular evaluation, the absence of follow-up outcome data prevents determination of whether symptom differences reflect cardiovascular pathology prevalence, environmental factors, or other mechanisms. These findings highlight important healthcare equity considerations, as increased symptom-driven evaluations place additional burden on already under-resourced populations. Future prospective research with comprehensive outcome assessment is needed to understand the clinical significance of these symptom patterns and develop equitable screening strategies.

## Data Availability

The deidentified participant data, data dictionary, and statistical analysis code supporting the findings of this study are available from the corresponding author upon reasonable request and execution of an appropriate data use agreement.

## List of Abbreviations

SCA: Sudden cardiac arrest
SCD: Sudden cardiac death
SDOH: Social determinants of health
COI: Child Opportunity Index
HCM: Hypertrophic cardiomyopathy AED: Automated external defibrillator
CPR: Cardiopulmonary resuscitation AHA: American Heart Association
ECG: Electrocardiogram
ADHD: Attention-deficit/hyperactivity disorder
LQTS: Long QT syndrome
ALCA-R: Anomalous left coronary artery from the right sinus of Valsalva

## References

1. Holmberg MJ, Ross CE, Fitzmaurice GM, et al. Annual Incidence of Adult and Pediatric In-Hospital Cardiac Arrest in the United States. Circ Cardiovasc Qual Outcomes. Jul 9 2019;12(7):e005580.

2. El-Assaad I, Al-Kindi SG, Aziz PF. Trends of Out-of-Hospital Sudden Cardiac Death Among Children and Young Adults. Pediatrics. Dec 2017;140(6)doi:10.1542/peds.2017-1438

3. Reinier K, Thomas E, Andrusiek DL, et al. Socioeconomic status and incidence of sudden cardiac arrest. Cmaj. Oct 18 2011;183(15):1705–12. doi:10.1503/cmaj.101512

4. Bernardin ME, Arora J, Schuler P, et al. Social determinants of health and their associations with outcomes in pediatric out-of-hospital cardiac arrest: A national study of the NEMSIS database. Resusc Plus. Dec 2024;20:100795. doi:10.1016/j.resplu.2024.100795

5. Kienbacher CL, Wei G, Rhodes J, Herkner H, Williams KA. Socioeconomic Risk Factors for Pediatric Out-of-hospital Cardiac Arrest: A Statewide Analysis. West J Emerg Med. Apr 28 2023;24(3):572–578. doi:10.5811/westjem.59107

6. Maron BJ, Friedman RA, Kligfield P, et al. Assessment of the 12-lead electrocardiogram as a screening test for detection of cardiovascular disease in healthy general populations of young people (12-25 years of age): a scientific statement from the American Heart Association and the American College of Cardiology. J Am Coll Cardiol. Oct 7 2014;64(14):1479–514. doi:10.1016/j.jacc.2014.05.006

7. Kim JH, Baggish AL, Levine BD, et al. Clinical Considerations for Competitive Sports Participation for Athletes With Cardiovascular Abnormalities: A Scientific Statement From the American Heart Association and American College of Cardiology. J Am Coll Cardiol. Mar 18 2025;85(10):1059–1108. doi:10.1016/j.jacc.2024.12.025

8. Maron BJ, Thompson PD, Ackerman MJ, et al. Recommendations and considerations related to preparticipation screening for cardiovascular abnormalities in competitive athletes: 2007 update: a scientific statement from the American Heart Association Council on Nutrition, Physical Activity, and Metabolism: endorsed by the American College of Cardiology Foundation. Circulation. Mar 27 2007;115(12):1643–455. doi:10.1161/circulationaha.107.181423

9. Shen WK, Sheldon RS, Benditt DG, et al. 2017 ACC/AHA/HRS Guideline for the Evaluation and Management of Patients With Syncope: A Report of the American College of Cardiology/American Heart Association Task Force on Clinical Practice Guidelines and the Heart Rhythm Society. Circulation. Aug 1 2017;136(5):e60–e122. doi:10.1161/cir.0000000000000499

10. Zipes DP, Link MS, Ackerman MJ, Kovacs RJ, Myerburg RJ, Estes NAM, 3rd. Eligibility and Disqualification Recommendations for Competitive Athletes With Cardiovascular Abnormalities: Task Force 9: Arrhythmias and Conduction Defects: A Scientific Statement From the American Heart Association and American College of Cardiology. J Am Coll Cardiol. Dec 1 2015;66(21):2412–2423. doi:10.1016/j.jacc.2015.09.041

11. Strickberger SA, Benson DW, Biaggioni I, et al. AHA/ACCF Scientific Statement on the evaluation of syncope: from the American Heart Association Councils on Clinical Cardiology, Cardiovascular Nursing, Cardiovascular Disease in the Young, and Stroke, and the Quality of Care and Outcomes Research Interdisciplinary Working Group; and the American College of Cardiology Foundation: in collaboration with the Heart Rhythm Society: endorsed by the American Autonomic Society. Circulation. Jan 17 2006;113(2):316–27. doi:10.1161/circulationaha.105.170274

12. Dalal A, Czosek RJ, Kovach J, et al. Clinical Presentation of Pediatric Patients at Risk for Sudden Cardiac Arrest. J Pediatr. Oct 2016;177:191–196. doi:10.1016/j.jpeds.2016.06.088

13. Bazoukis G, Loscalzo J, Hall JL, Bollepalli SC, Singh JP, Armoundas AA. Impact of Social Determinants of Health on Cardiovascular Disease. J Am Heart Assoc. Mar 4 2025;14(5):e039031. doi:10.1161/jaha.124.039031

14. Bevan GH, Nasir K, Rajagopalan S, Al-Kindi S. Socioeconomic Deprivation and Premature Cardiovascular Mortality in the United States. Mayo Clin Proc. Jun 2022;97(6):1108–1113. doi:10.1016/j.mayocp.2022.01.018

15. Arthur MN, DeLong RN, Kucera K, et al. Socioeconomic deprivation and racialised disparities in competitive athletes with sudden cardiac arrest from the USA. Br J Sports Med. Apr 25 2024;58(9):494–499. doi:10.1136/bjsports-2023-107367

16. Noelke, C., McArdle, N., DeVoe, B., Leonardos, M., Lu, Y., Ressler, R.W., & Acevedo-Garcia, D. (2024). Child Opportunity Index 3.0 Technical Documentation. diversitydatakids.org, Brandeis University. Retrieved from diversitydatakids.org/research-library/coi-30-technical-documentation.

17. Corsi D, Saraiya A, Doyle M, et al. Cardiac screening findings and referral patterns in male African-American basketball players: Analysis of the HeartBytes Registry. Am J Cardiol. May 15 2025;243:73–80. doi:10.1016/j.amjcard.2025.02.007

18. Al-Khatib SM, Stevenson WG, Ackerman MJ, et al. 2017 AHA/ACC/HRS guideline for management of patients with ventricular arrhythmias and the prevention of sudden cardiac death: Executive summary: A Report of the American College of Cardiology/American Heart Association Task Force on Clinical Practice Guidelines and the Heart Rhythm Society. Heart Rhythm. Oct 2018;15(10):e190-e252. doi:10.1016/j.hrthm.2017.10.035

19. Arnett DK, Blumenthal RS, Albert MA, et al. 2019 ACC/AHA Guideline on the Primary Prevention of Cardiovascular Disease: A Report of the American College of Cardiology/American Heart Association Task Force on Clinical Practice Guidelines. Circulation. Sep 10 2019;140(11):e596-e646. doi:10.1161/cir.0000000000000678

20. Weintraub WS, Daniels SR, Burke LE, et al. Value of primordial and primary prevention for cardiovascular disease: a policy statement from the American Heart Association. Circulation. Aug 23 2011;124(8):967–90. doi:10.1161/CIR.0b013e3182285a81

21. Baggish AL, Battle RW, Beaver TA, et al. Recommendations on the Use of Multimodality Cardiovascular Imaging in Young Adult Competitive Athletes: A Report from the American Society of Echocardiography in Collaboration with the Society of Cardiovascular Computed Tomography and the Society for Cardiovascular Magnetic Resonance. J Am Soc Echocardiogr. May 2020;33(5):523–549. doi:10.1016/j.echo.2020.02.009

22. Baggish AL, Battle RW, Beckerman JG, et al. Sports Cardiology: Core Curriculum for Providing Cardiovascular Care to Competitive Athletes and Highly Active People. J Am Coll Cardiol. Oct 10 2017;70(15):1902–1918. doi:10.1016/j.jacc.2017.08.055

23. Zhang C, Shi D. Left anomalous coronary artery originating from the opposite sinus causes acute myocardial infarction with syncope in a young woman: A case report. Medicine (Baltimore). Sep 27 2024;103(39):e39850. doi:10.1097/md.0000000000039850

24. Palmieri V, Gervasi S, Bianco M, et al. Anomalous origin of coronary arteries from the "wrong" sinus in athletes: Diagnosis and management strategies. Int J Cardiol. Feb 1 2018;252:13–20. doi:10.1016/j.ijcard.2017.10.117

25. Erickson CC, Salerno JC, Berger S, et al. Sudden Death in the Young: Information for the Primary Care Provider. Pediatrics. Jul 2021;148(1)doi:10.1542/peds.2021-052044

26. Newman DB, Garmany R, Contreras AM, et al. Cardiopulmonary Exercise Testing in Athletes With Hypertrophic Cardiomyopathy. Am J Cardiol. Feb 15 2023;189:49–55. doi:10.1016/j.amjcard.2022.11.008

27. Blaustein JR, Quisel MJ, Hamburg NM, Wittkopp S. Environmental Impacts on Cardiovascular Health and Biology: An Overview. Circ Res. Apr 26 2024;134(9):1048–1060. doi:10.1161/circresaha.123.323613

28. Cutrufello PT, Smoliga JM, Rundell KW. Small things make a big difference: particulate matter and exercise. Sports Med. Dec 1 2012;42(12):1041–58. doi:10.1007/bf03262311

29. Goossens J, Jonckheere AC, Seys SF, et al. Activation of epithelial and inflammatory pathways in adolescent elite athletes exposed to intense exercise and air pollution. Thorax. Aug 2023;78(8):775–783. doi:10.1136/thorax-2022-219651

30. Vine M, Vander Stoep A, Bell J, Rhew IC, Gudmundsen G, McCauley E. Associations between household and neighborhood income and anxiety symptoms in young adolescents. Depress Anxiety. Sep 2012;29(9):824–32. doi:10.1002/da.21948

31. Kowalchuk A, Gonzalez SJ, Zoorob RJ. Anxiety Disorders in Children and Adolescents. Am Fam Physician. Dec 2022;106(6):657–664.

32. Agarwala A, Patel J, Stephens J, et al. Implementation of Prevention Science to Eliminate Health Care Inequities in Achieving Cardiovascular Health: A Scientific Statement From the American Heart Association. Circulation. Oct 10 2023;148(15):1183–1193. doi:10.1161/cir.0000000000001171

33. Magnani JW, Mujahid MS, Aronow HD, et al. Health Literacy and Cardiovascular Disease: Fundamental Relevance to Primary and Secondary Prevention: A Scientific Statement From the American Heart Association. Circulation. Jul 10 2018;138(2):e48–e74. doi:10.1161/cir.0000000000000579

34. Al-Khatib SM, Yancy CW, Solis P, et al. 2016 AHA/ACC Clinical Performance and Quality Measures for Prevention of Sudden Cardiac Death: A Report of the American College of Cardiology/American Heart Association Task Force on Performance Measures. J Am Coll Cardiol. Feb 14 2017;69(6):712–744. doi:10.1016/j.jacc.2016.09.933

35. Weisfeldt ML, Sitlani CM, Ornato JP, et al. Survival after application of automatic external defibrillators before arrival of the emergency medical system: evaluation in the resuscitation outcomes consortium population of 21 million. J Am Coll Cardiol. Apr 20 2010;55(16):1713–20. doi:10.1016/j.jacc.2009.11.077

36. Leung KHB, Brooks SC, Clegg GR, Chan TCY. Socioeconomically equitable public defibrillator placement using mathematical optimization. Resuscitation. Sep 2021;166:14–20. doi:10.1016/j.resuscitation.2021.07.002

37. van Hattum JC, Verwijs SM, Senden PJ, et al. The Sports Cardiology Team: Personalizing Athlete Care Through a Comprehensive, Multidisciplinary Approach. Mayo Clin Proc Innov Qual Outcomes. Dec 2022;6(6):525–535. doi:10.1016/j.mayocpiqo.2022.08.006

